# Excess death among Latino people in California during the COVID-19 pandemic

**DOI:** 10.1101/2020.12.18.20248434

**Authors:** Alicia R. Riley, Yea-Hung Chen, Ellicott C. Matthay, M. Maria Glymour, Jacqueline M. Torres, Alicia Fernandez, Kirsten Bibbins-Domingo

**Author notes:** Corresponding author: Dr. Kirsten Bibbins-Domingo, Professor and Chair of Department of Epidemiology and Biostatistics, UCSF, 550 16^th^ Street, San Francisco, CA 94158.

## Abstract

**Background:** Latino people in the US are experiencing higher excess deaths during the COVID-19 pandemic than any other racial/ethnic group, but it is unclear which subgroups within this diverse population are most affected. Such information is necessary to target policies that prevent further excess mortality and reduce inequities.

**Methods:** Using death certificate data for January 1, 2016 through February 29, 2020 and time-series models, we estimated the expected weekly deaths among Latino people in California from March 1 through October 3, 2020. We quantified excess mortality as observed minus expected deaths and risk ratios (RR) as the ratio of observed to expected deaths. We considered subgroups defined by age, sex, place of birth, education, occupation, and combinations of these factors.

**Findings:** During the first seven months of the pandemic, Latino deaths in California exceeded expected deaths by 10,316, a 31% increase. Excess death rates were greatest for individuals born in Mexico (RR 1.44; 95% PI, 1.41, 1.48) or Central America (RR 1.49; 95% PI, 1.37, 1.64), with less than a high school degree (RR 1.41; 95% PI, 1.35, 1.46), or in food-and-agriculture (RR 1.60; 95% PI, 1.48, 1.74) or manufacturing occupations (RR 1.59; 95% PI, 1.50, 1.69). Immigrant disadvantages in excess death were magnified among working-age Latinos in essential occupations.

**Interpretation:** The pandemic has disproportionately impacted mortality among Latino immigrants and Latinos in unprotected essential jobs; Interventions to reduce these disparities should include early vaccination, workplace safety enforcement, and expanded access to medical care.

**Funding:** National Institute on Aging; UCSF

**RESEARCH IN CONTEXT:** *Evidence before this study:* Several articles have suggested all-cause excess mortality estimates are superior to official COVID-19 counts for assessing the impact of the pandemic on marginalized populations that lack access to testing and healthcare. We searched PubMed, Google scholar, and the medRxiv preprint database through December 22, 2020 for studies of (“excess mortality” or “excess death”) AND (“COVID-19” or “coronavirus”) set in the United States and we identified two empirical studies with estimates of excess mortality among Latinos during the pandemic. The study set in California (from our research team) found per capita excess mortality was highest among Black and Latino people. The national study found percent excess mortality was significantly higher among Latino people than any other racial/ethnic group. Neither study further disaggregated the diverse Latino population or provided subgroup estimates to clarify why excess pandemic mortality is so high in this population. In the U.S., official COVID-19 statistics are rarely disaggregated by place of birth, education, or occupation which has resulted in a lack of evidence of how these factors have impacted mortality during the pandemic. No study to date of excess mortality in the U.S. has provided estimates for immigrant or occupational subgroups.

*Added value of this study:* Our population-based observational study of all-cause mortality during the COVID-19 pandemic provides the first estimates of within-group heterogeneity among the Latino population in California – one of the populations hardest hit by COVID-19 in the U.S. We provide the first subgroup estimates by place of birth and occupational sector, in addition to combined estimates by foreign-birth and participation in an essential job and education. In doing so, we reveal that Latino immigrants in essential occupations have the highest risk of excess death during the pandemic among working-age Latinos. We highlight the heightened risk of excess mortality associated with food/agriculture and manufacturing occupational sectors, essential sectors in which workers may lack COVID-19 protections.

*Implications of all the available evidence:* Our study revealed stark disparities in excess mortality during the COVID-19 pandemic among Latinos, pointing to the particularly high vulnerability of Latino immigrants and Latinos in essential jobs. These findings may offer insight into the disproportionate COVID-19 mortality experienced by immigrants or similarly marginalized groups in other contexts. Interventions to reduce these disparities should include policies enforcing occupational safety, especially for immigrant workers, early vaccination, and expanded access to medical care.

## INTRODUCTION

Throughout the coronavirus disease 2019 (COVID-19) pandemic, Latino people in the US have been dying at disproportionately high rates. National all-cause mortality increased 53.6% for Latinos from the start of the pandemic through October – more than any other group and over four times more than the percent increase for non-Latino Whites^1^. However, aggregate national and state-level statistics obscure the vast diversity among Latinos in the US, making it difficult to intervene effectively to prevent further pandemic mortality^2^. Identifying subgroups at greatest risk for excess death could help guide targeted interventions including workplace protections, testing, and vaccination.

The state of California is a critical setting to evaluate the drivers of Latino mortality during the COVID-19 pandemic. California is home to approximately one quarter of the U.S.’s Latino population and its immigrant population. Latinos are 39% of California’s population of 39.5 million residents, but they account for 48% of COVID-19-confirmed deaths through December 2020^3^. Even this remarkable death toll likely understates the impact of the pandemic on Latinos because COVID-19-confirmed deaths are understood to be a subset of the total deaths due to COVID-19^4^.

The current study estimates the impact of the COVID-19 pandemic on deaths among Latino individuals in California by estimating excess deaths that occurred between March 1, 2020 and October 3, 2020 compared to the four years prior to the COVID-19 pandemic. By examining death record data, this study overcomes the limitations of official COVID-19 death counts^5^. In addition, with the overarching goal of assisting public health efforts globally, we evaluate heterogeneous risk within the Latino population by examining excess death during the pandemic by age, sex, place of birth, education, and occupation.

## METHODS

### Data

In order to forecast expected mortality trends during our time-period of interest, we drew on death certificate data from the California Department of Public Health for the period from January 3, 2016 to February 29, 2020 (pre-pandemic period) and from March 1, 2020 to October 3, 2020 (pandemic period). Our analyses included all deaths that occurred within the state of California for which Hispanic/Latino ethnicity was designated. (Only 0.29% of deaths during the pandemic period and 0.25% of deaths during the pre-pandemic period lacked information on ethnicity). Our analytic sample consisted of 43,576 total deaths among Latinos in California during the pandemic period and 220,986 during the pre-pandemic period. There was little missingness for the variables of interest and exclusions due to missingness were done on an available-case basis. Due to delays in complete cause of death data, we did not distinguish specific causes of death for this analysis.

We defined Latinos as any individual identified with Latino/a or Hispanic ethnicity on the death certificate, irrespective of race or country of birth. We do not use the more inclusive, gender-neutral terms Latinx or Latine in this paper because they were not used in death records.

We evaluated subgroups defined by: age at death in years (0-24, 25-54, 55-64, 65-74, 75-84, or 85+); sex (male or female); place of birth (4-categories: U.S., Mexico, Central America, or other; 2-categories: U.S., or foreign-born); occupational sector (see below); educational attainment (4-categories: no high school degree, high school degree or GED, some college or Associate’s degree, Bachelor’s degree or beyond; 2-categories: no high school degree, or high school or more).

### Outcome

We defined excess deaths as observed minus expected deaths due to all causes, with expected deaths based on historical trends over the previous four years using time series methods described below. For analyses stratified by educational attainment, following standard practice, we restricted our analysis to decedents at least 25 years of age so most people would have completed schooling. For analyses stratified by occupational sector, we grouped individuals by the primary occupation over their life, a field included in the death certificate, and restricted analyses to decedents 18 to 65 years of age. We drew on the California Department of Public Health designations for essential work^6^ to sort the occupation into one of seven sectors of high-risk or essential work, a category for non-essential, or a category for unemployed or missing data on occupation. Using this designation, we also compared individuals in essential occupational sectors to individuals in non-essential occupational sectors.

### Statistical Analyses

For each subgroup of interest, we repeated the following procedure using the forecast package in R^7^. First, we aggregated the data to weekly death counts. Next, to estimate expected deaths, we fit dynamic harmonic regression models with autoregressive integrated moving average (ARIMA)^8^ errors for the number of weekly deaths, using deaths occurring pre-pandemic. We iterated through models with up to 25 Fourier terms and selected the model that minimized the corrected Akaike information criterion, following an approach developed by Hyndman and Khandakar^7^. We inspected the residuals from the preferred model using plots and the Ljung-Box test to confirm that minimal autocorrelation remained. Then, using the preferred model, we forecast the number of weekly deaths and the corresponding 95% prediction intervals (PI) for the pandemic period: March 1, 2020 through October 3, 2020. These forecasts provided the expected weekly deaths during the pandemic period based on historical trends. To estimate the weekly excess deaths, we subtracted the number of expected deaths from the observed deaths for each week. We constructed a 95% PI for excess deaths by simulating the expected deaths model 10,000 times (varying based on uncertainty in the estimated coefficients for the time-series prediction model), selecting the 2.5 and 97.5 percentiles, and subtracting the total number of observed deaths. Risk ratios were estimated as observed deaths divided by expected deaths, with the pandemic as the exposure, and are also known as observed-to-expected mortality ratios. Risk ratios present the same information as percent excess deaths, as reported in previous studies^1,9,9^. We also calculated per capita excess mortality by dividing excess deaths by the corresponding population size, using estimates from the 2019 American Community Survey. We conducted all analyses in R version 3.6.3.

## RESULTS

Between March 1 and October 3, 2020, Latino people in California experienced a 31% increase in mortality compared with historical trends, with an estimated 10,316 (95% PI, 9,570, 11,030) excess deaths (Table 1). COVID-19 was listed as a cause of death on 7672 death certificates (74% of the estimated excess deaths). The number of excess deaths varied by week such that excess mortality was lowest during California’s shelter-in-place period (March 19, 2020-May 7, 2020), then increased to a peak at the end of July with over 600 excess deaths per week, and began to decline by mid-August (Figure 3), but the time trends varied by subpopulation (Figure 4). The observed excess death rate for Latinos aged 25 or older implies 1185 additional deaths per million individuals. Excess deaths in Latinos were observed in all regions in California (Figure S1).

**Table 1.** Cumulative excess deaths during the COVID-19 pandemic among Latinos in California: March through September 2020.

|  | Excess | Risk ratio* | Per 100,000 |
| --- | --- | --- | --- |
| Entire state | 10316 (9570, 11030) | 1.31 (1.28, 1.34) | 66 |
| Age |  |  |  |
| 0-24 years old | 184 (-17, 376) | 1.10 (0.99, 1.23) | 3 |
| 25-54 years old | 2346 (1965, 2708) | 1.38 (1.30, 1.46) | 36 |
| 55-64 years old | 1986 (1689, 2271) | 1.40 (1.32, 1.48) | 146 |
| 65-74 years old | 2425 (2018, 2816) | 1.42 (1.33, 1.52) | 325 |
| 75-84 years old | 2110 (1809, 2398) | 1.35 (1.28, 1.41) | 641 |
| 85+ years old | 1978 (1472, 2464) | 1.26 (1.18, 1.35) | 1535 |
| Gender |  |  |  |
| Female | 3745 (3101, 4360) | 1.25 (1.20, 1.31) | 48 |
| Male | 7092 (6214, 7937) | 1.39 (1.33, 1.46) | 90 |
| Country of birth |  |  |  |
| United States | 3004 (2506, 3482) | 1.19 (1.16, 1.23) | 29 |
| Mexico | 5855 (5497, 6199) | 1.44 (1.41, 1.48) | 149 |
| Central America | 1311 (1068, 1543) | 1.49 (1.37, 1.64) | 144 |
| Other | 223 (91, 350) | 1.15 (1.06, 1.27) | 76 |
| Education† |  |  |  |
| No high school degree and no GED | 5936 (5375, 6471) | 1.41 (1.35, 1.46) | 193 |
| High school degree or GED | 2766 (2233, 3279) | 1.31 (1.23, 1.39) | 114 |
| Some college or associate degree | 1038 (719, 1344) | 1.23 (1.15, 1.32) | 44 |
| Bachelor's degree or beyond | 435 (245, 616) | 1.22 (1.11, 1.34) | 33 |
| Occupational Sector‡ |  |  |  |
| Facilities | 883 (697, 1063) | 1.38 (1.28, 1.50) | 58 |
| Food and agriculture | 639 (551, 723) | 1.60 (1.48, 1.74) | 61 |
| Government and community | 249 (208, 288) | 1.42 (1.33, 1.53) | 33 |
| Health or emergency | 202 (128, 273) | 1.35 (1.20, 1.54) | 35 |
| Manufacturing | 519 (465, 573) | 1.59 (1.50, 1.69) | 101 |
| Retail | 244 (168, 318) | 1.29 (1.18, 1.41) | 35 |
| Transportation and logistics | 804 (660, 942) | 1.39 (1.30, 1.49) | 85 |
| Not essential | 1119 (921, 1310) | 1.30 (1.23, 1.36) | 55 |
| Unemployed or missing | 167 (96, 236) | 1.40 (1.19, 1.66) | 10 |

\*: Risk ratio is observed/expected for all-cause mortality, comparing the pandemic period (March 1, 2020 to October 3, 2020) to previous years (January 3, 2016 to February 29, 2020).
†: Among decedents 25 years of age or older.
‡: Among decedents 18–65 years of age.
Sources: California Department of Public Health Death Records; 2019 American Community Survey.

**Figure 1.**
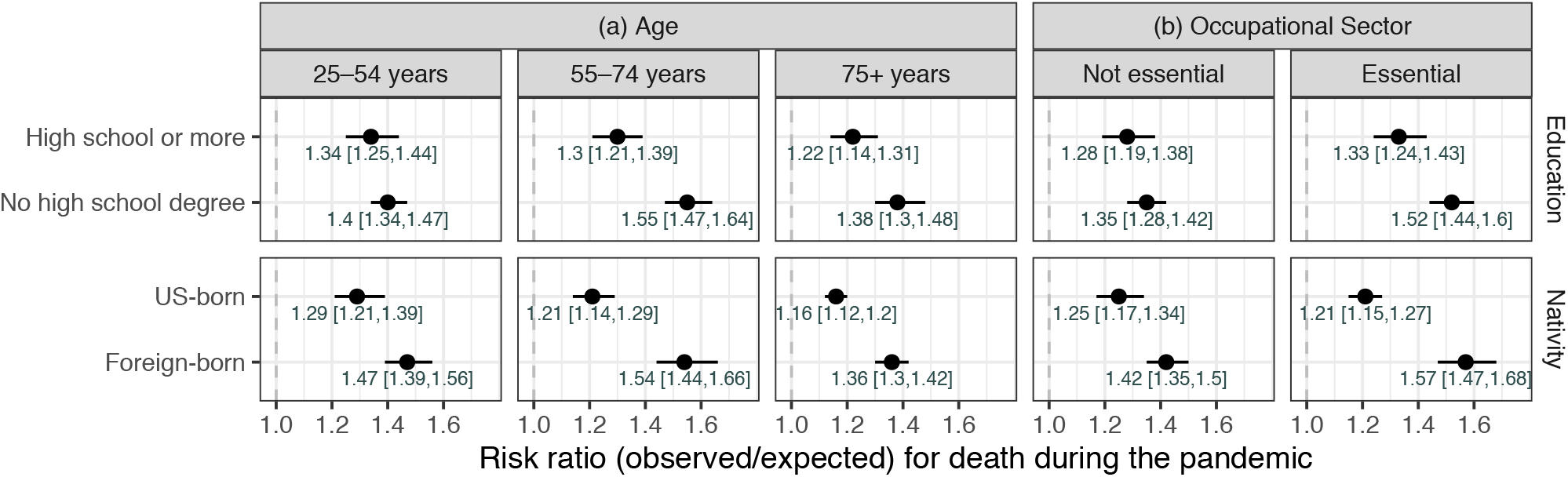
Cumulative excess mortality during the COVID-19 pandemic among Latinos in California, by subgroups. a: Among decedents 25 years of age at death or older. b: Among decedents 25 to 65 years of age at death. Note: Risk ratio is observed/expected for all-cause mortality, comparing the pandemic period (March 1, 2020 to October 3, 2020) to previous years (January 3, 2016 to February 29, 2020); Source: California Department of Public Health Death Records.

**Figure 2.**
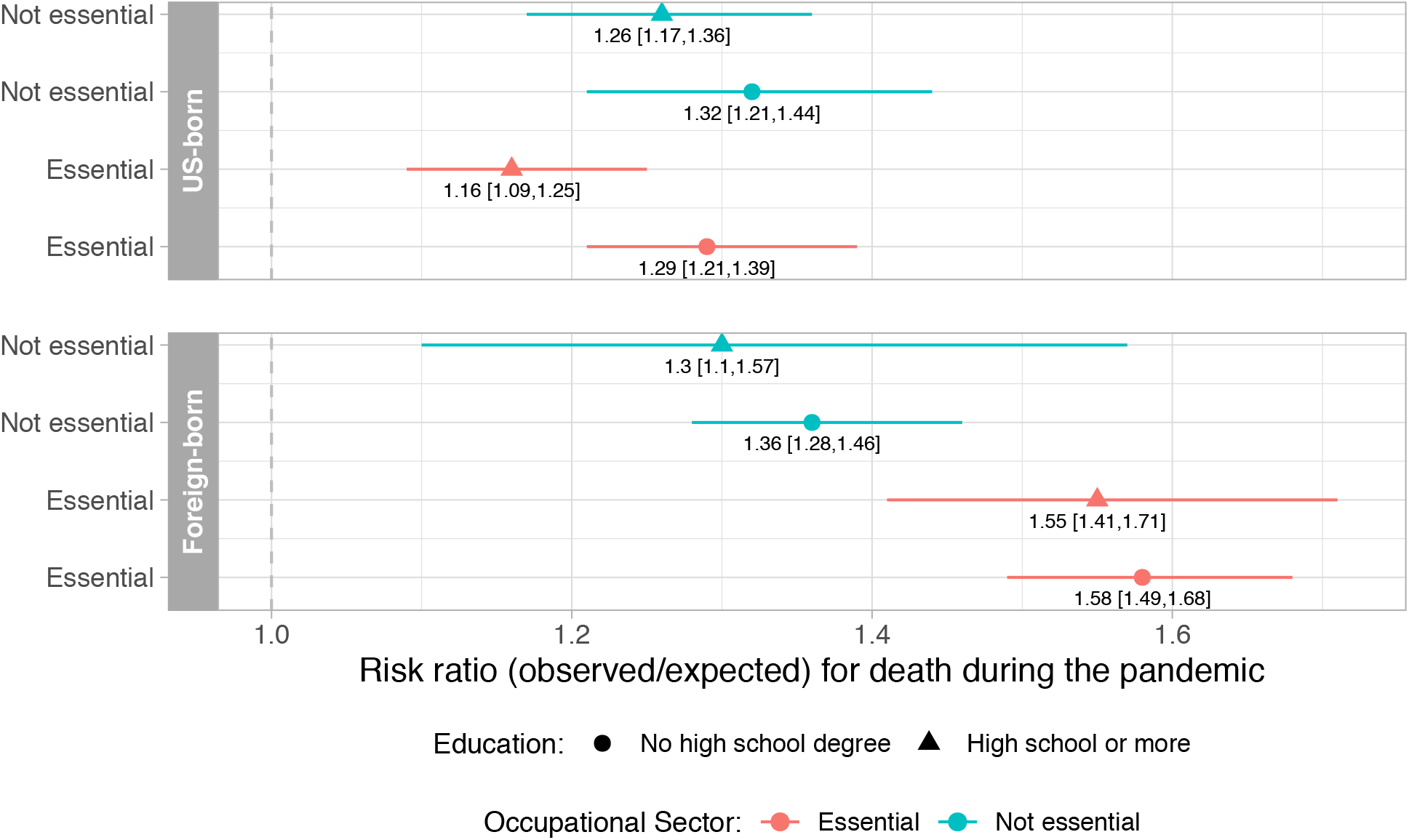
Cumulative excess mortality during the COVID-19 pandemic among Latinos in California, by intersecting subgroups (nativity by education by occupational sector) Note: Analysis restricted to decedents 25 to 65 years of age at death. Risk ratio is observed/expected for all-cause mortality, comparing the pandemic period (March 1, 2020 to October 3, 2020) to previous years (January 3, 2016 to February 29, 2020); Source: California Department of Public Health Death Records.

**Figure 3.**
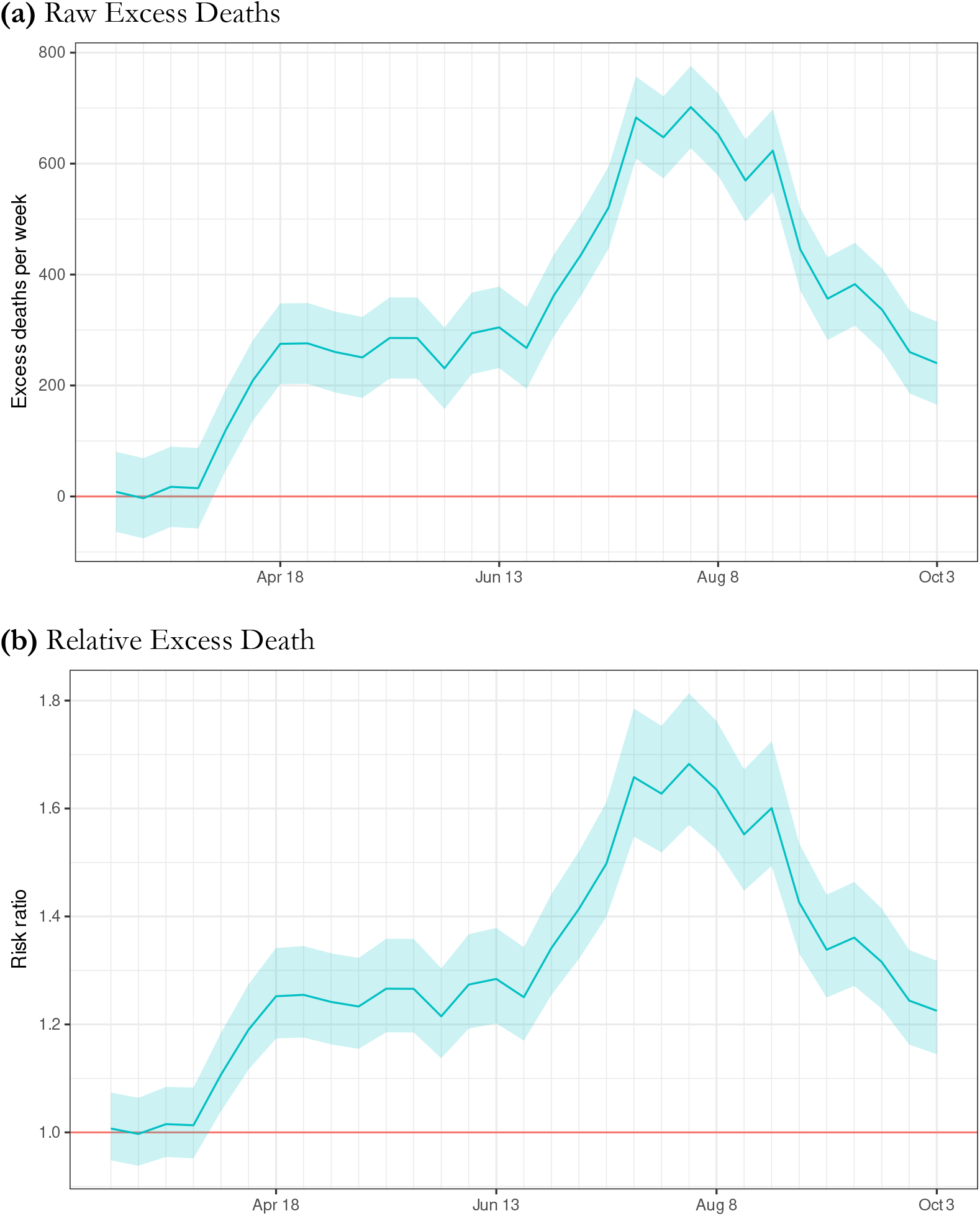
Weekly excess mortality during the COVID-19 pandemic among Latinos in California, March through September 2020. Note: Risk ratio is observed/expected for all-cause mortality, comparing the pandemic period (March 1, 2020 to October 3, 2020) to previous years (January 3, 2016 to February 29, 2020); Source: California Department of Public Health Death Records.

**Figure 4.**
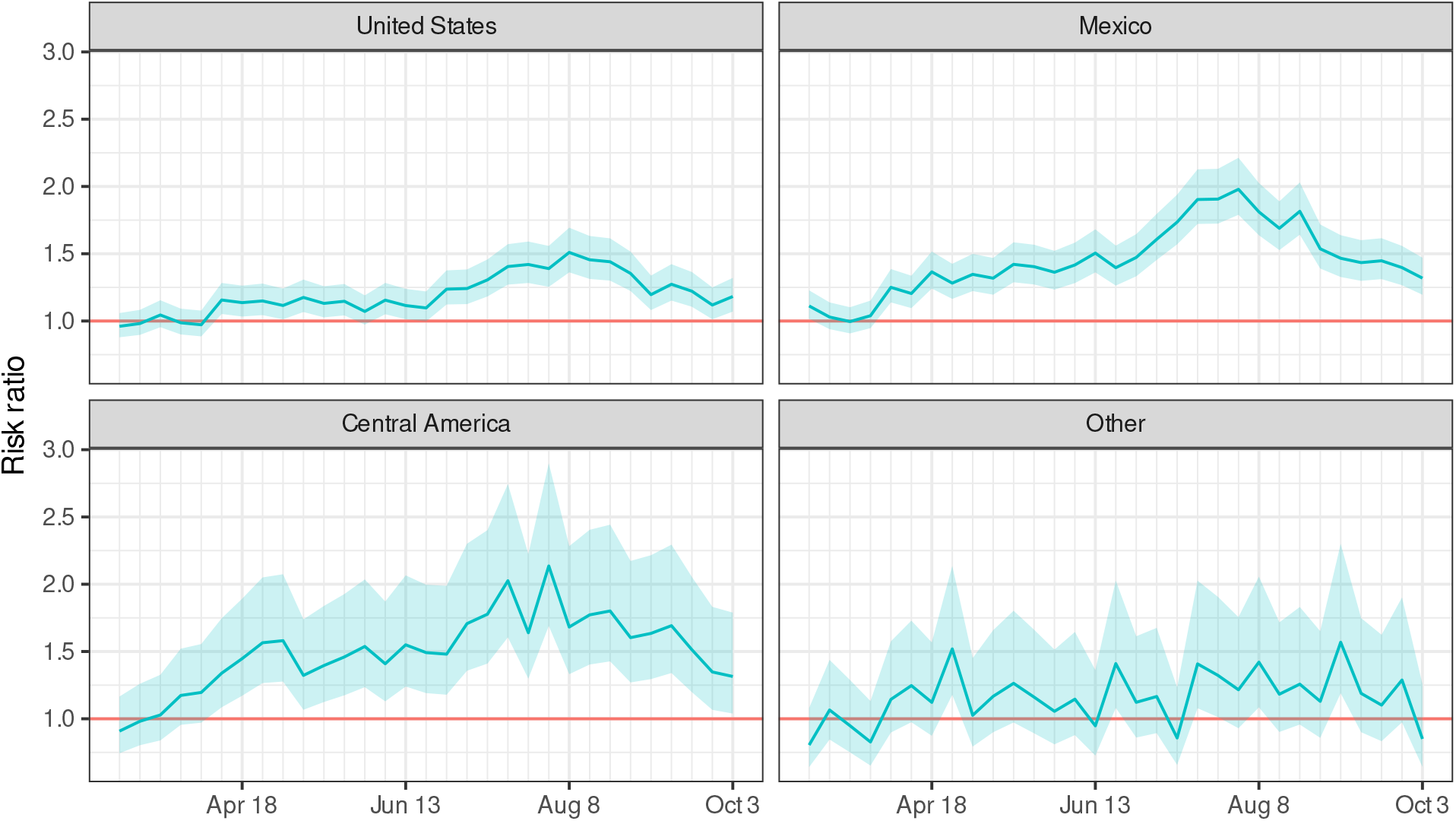
Weekly excess mortality during the COVID-19 pandemic among Latinos in California, by place of birth. Note: Risk ratio (observed/expected) for all-cause mortality, comparing the pandemic period (March 1, 2020 to October 3, 2020) to previous years (January 3, 2016 to February 29, 2020); Source: California Department of Public Health Death Records.

As shown in Table 1, relative increases in mortality ranged from 26-42% for all adults 25 and older and was of similar magnitude across adult age groups. The magnitude of excess mortality varied by country of birth and education. Foreign-born Latinos had higher relative excess mortality than U.S.-born Latinos: deaths were 44 percent higher than expected among individuals born in Mexico (RR 95% PI, 1.41, 1.48) and 49 percent higher than expected among individuals born in Central America (RR 95% PI, 1.37, 1.64). By contrast, deaths among U.S.-born Latinos were 19 percent (RR 95% PI, 1.16, 1.23) higher than expected values. In every adult age stratum, foreign-born Latinos experienced at least double the relative excess mortality experienced by U.S.-born Latinos (Figure 1). Educational attainment was inversely associated with the magnitude of excess death during the pandemic period. Relative excess mortality was highest among Latinos with less than a high school degree (RR 1.41; 95% PI, 1.35, 1.46) and lowest among Latinos with a Bachelor’s degree or more (RR 1.22; 95% PI, 1.11, 1.34).

Among working-age adults (age 18 to 65 years), cumulative excess mortality RRs were highest in those working in food-and-agriculture (RR 1.60; 95% PI, 1.48, 1.74) and manufacturing (RR 1.59; 95% PI, 1.50, 1.69) (Table 1). During the summer surge, weekly excess mortality among workers in manufacturing was over 200 percent of expected values (Figure 4).

Disadvantages in excess mortality associated with foreign-birth and low education were more pronounced among Latinos in essential occupations and in the 55 to 74 age group. Among Latinos who worked in essential sectors, the percent excess in mortality for immigrants (RR 1.57; 95% PI, 1.47, 1.68) was more than double the percent excess for U.S.-born Latinos (RR 1.21; 95% PI, 1.15, 1.27) (Figure 1). This 36-percentage-point immigrant disadvantage was reduced to 17-percentage-points among Latinos in non-essential occupations. Among Latinos in essential occupational sectors who lacked a high school credential, mortality was 52% (95% PI, 44%, 60%) higher than expected, whereas for Latinos in the same occupational sectors with a high school credential or more, mortality was 33% (95% PI, 24%, 43%) higher than expected. Finally, pandemic mortality was 58 percent (95% PI, 49%, 68%) above expected in the subgroup of working-age Latinos with the combined factors of being foreign-born, without a high school degree, and in an essential occupation, but significantly lower in the subgroup of Latinos who were foreign-born and without a high school degree, yet not in an essential occupation (RR 1.36; 95% PI, 1.28, 1.48) (Figure 2).

## DISCUSSION

Using data on all deaths among Latinos in California, USA from January 1, 2016 through October 3, 2020, we found 10,316 additional deaths occurred among California Latinos in the first 7 months of the pandemic compared with trends in the four years prior. In relative terms, this was an excess of 31 percent. Our findings reveal disparities in excess mortality by Latino subgroups. Immigrant Latinos, those with less than a high school degree, and those working in essential occupations, in particular food/agriculture and manufacturing, were at markedly higher risk of death, especially in the presence of more than one of these risk factors. Participation in an essential occupational sector appeared to magnify existing social vulnerabilities.

We have previously shown that over the first 5 months of the pandemic, Latinos in California experienced higher excess mortality than other racial/ethnic groups, with a per capita excess nearly double that of non-Latino White people in California^10^. Our current results show that the pandemic’s impacts on mortality have been far worse for Central American-born and Mexican-born Latinos than U.S.-born Latinos; a disparity apparent within every age stratum, but worse among 55 to 74 year-olds. Latino immigrants comprise 37% of California’s Latino population, but an estimated 71 percent of Latino excess deaths during the pandemic period occurred among immigrants.

The differential impact on foreign-born Latinos is notable given previously documented immigrant health advantages^11^, and may have relevance to other countries. The literature suggests recent Latino immigrants to the U.S. are less likely than their U.S.-born counterparts to have an underlying medical condition that would increase COVID-19 severity^12^. However, foreign-born Latinos may be more vulnerable to COVID-19 because of their concentration in high-exposure occupations with limited workplace protections^13^, barriers to health care and other social protections due to legal status and public charge inadmissibility^14^, and chilling effects on healthcare-seeking behavior due to anti-immigrant rhetoric and policy^15^.

The elevated mortality and stark disparities observed in essential occupational sectors underscores the potential role of workplace settings and worker marginalization as contributors to excess deaths among Latino individuals^16,17^, and as shown in Figure 2, particularly among immigrants. The risk ratios for excess death among those in health and emergency jobs or government and community jobs – where workplace safety measures are more consistently adopted^18^ --were substantially lower than the risk ratios among those in sectors characterized by low-wage jobs such as manufacturing and agriculture. Notably, the foreign-born disadvantage and the low-education disadvantage in pandemic mortality were both magnified among Latinos in essential occupational sectors (Figure 1). Subgroup analyses revealed that essential work dramatically increased the risk of death among foreign-born Latinos, but not among U.S.-born Latinos (Figure 2), perhaps highlighting differences in the protections available to workers.

Multiple structural inequalities may put foreign-born and low-educated Latinos employed in essential jobs and their family members at greater risk of exposure to SARS-CoV-2 and death from COVID-19. For instance, foreign-born Latinos earn 87% as much as their U.S.-born counterparts^13^. Extreme financial precarity coupled with low levels of unionization make it difficult to negotiate workplace protections and benefits^19^, leading foreign-born and low-educated Latinos to participate in work settings where they are not protected from exposure. Also, approximately 44% of Mexican immigrants in the U.S.^20^ and 69% of Guatemalan immigrants in the U.S. are undocumented^21^. The threat of detention or deportation combined with exclusion from Medicaid and Medicare result in delayed and inadequate medical care for many undocumented Latinos^19,22^. Among Latinos in California, half of undocumented adults and over 20 percent of legal permanent resident adults lacked health insurance prior to the pandemic^23^. In addition to these structural factors, many Latinos for whom Spanish or an indigenous language is their preferred language may lack access to actionable health information about COVID-19 prevention and treatment^24^. Larger households and denser living arrangements likely amplify occupational exposures and contribute to the elevated case and death rates among Latinos^16^. Within these intersecting factors are many opportunities for intervention.

The two main strategies for pandemic control endorsed during the study period were individual behavior change (e.g., masking, distancing) and economic shutdowns to slow transmission. Our results suggest that neither pandemic control strategy has sufficiently protected Latinos from excess mortality. In fact, as our previous study demonstrated and our current study reinforces, California began reopening in May when excess deaths among Latinos had yet to decline and, as we show here, when excess death were rising for vulnerable subgroups. Distinct policies and investment are needed to protect immigrant Latinos and reduce disparities in COVID-19-related death^25–27^.

Our findings draw attention to four domains of promising policy response to reduce excess pandemic mortality among marginalized groups: workplace conditions, financial supports, healthcare coverage, and vaccine distribution. First, expanded action is needed, particularly in the manufacturing and food & agriculture sectors, to regulate and enforce workplace modifications to protect employees from SARS-CoV-2 exposure, provide workers with accurate information, and protect jobs if workers take sick leave or negotiate for safer conditions^28^. Second, most federal government programs in the U.S. to provide financial relief from the economic consequences of the pandemic have excluded undocumented individuals and the many Latinos in mixed status families. Including immigrant families in financial relief could enable more workers to refuse to work in unsafe settings, which might interrupt transmission and reduce mortality. Third, an immediate step that could improve access to medical care is extending emergency Medicaid coverage to all individuals with COVID-19 regardless of immigration status^29^. Finally, pandemic disparities are likely to widen in the U.S. and similar contexts unless action is taken to prioritize marginalized populations, including unauthorized immigrants and those without health insurance, for access to COVID-19 vaccination.

This study is limited by the accuracy and comprehensiveness of the data on death certificates. The interpretation of excess deaths that did not list COVID-19 as a contributing cause is unclear; these deaths may be undiagnosed COVID-19 cases or they may reflect increased mortality due to other causes exacerbated by the pandemic. The proportion of COVID-confirmed deaths compared with overall excess mortality that we observed (approximately 74%) is similar or slightly higher than in national studies. Our analysis is also limited by the occupation field on death certificates which indicates primary occupation for most of the decedent’s lifetime rather than most recent occupation. We did not have data to directly examine a number of plausible contributors to excess mortality among Latinos, for example household size or legal status. Nor did we have access to data on indigenous ethnicity, another potentially important subgroup with high vulnerability^30^. Finally, our results rest on the time-series model premise, that we can predict mortality rates in 2020 based on trends observed in the prior four years, accounting for factors such as seasonality and autocorrelation.

In summary, our study used data from individual death records to reveal that immigrant Latinos, Latinos with low education, and Latinos in essential jobs have experienced particularly high excess mortality during the COVID-19 pandemic. Policies enforcing occupational safety, especially for immigrant workers, and policies extending high-quality healthcare to all people, including immigrants, may have the highest benefit for reducing COVID-19 mortality.

## Data Availability

The data can be requested directly from the California Department of Public Health by filing a Vital Records Data Application available at this website: https://www.cdph.ca.gov/Programs/CHSI/Pages/Data-Applications.aspx

## Ethics statement

The study protocol was approved by the Committee for the Protection of Human Subjects (State of California), project number: 2020-109.

## Contributors

ARR, YC, and ECM had full access to the data and take responsibility for the analysis. Study design: All authors; Data analysis: Led by YC and reviewed by ARR and ECM; Data interpretation: All authors; First draft of manuscript: ARR; Revisions and approval of final draft of manuscript and tables/figures: All authors. Supervision: KBD.

## Data sharing

The death record data used in this study are restricted and were made available by the California Department of Public Health through a Data Use Agreement. To apply for access visit: https://www.cdph.ca.gov/Programs/CHSI/Pages/Data-Applications.aspx.

## Supplementary Material

**Figure S1.**
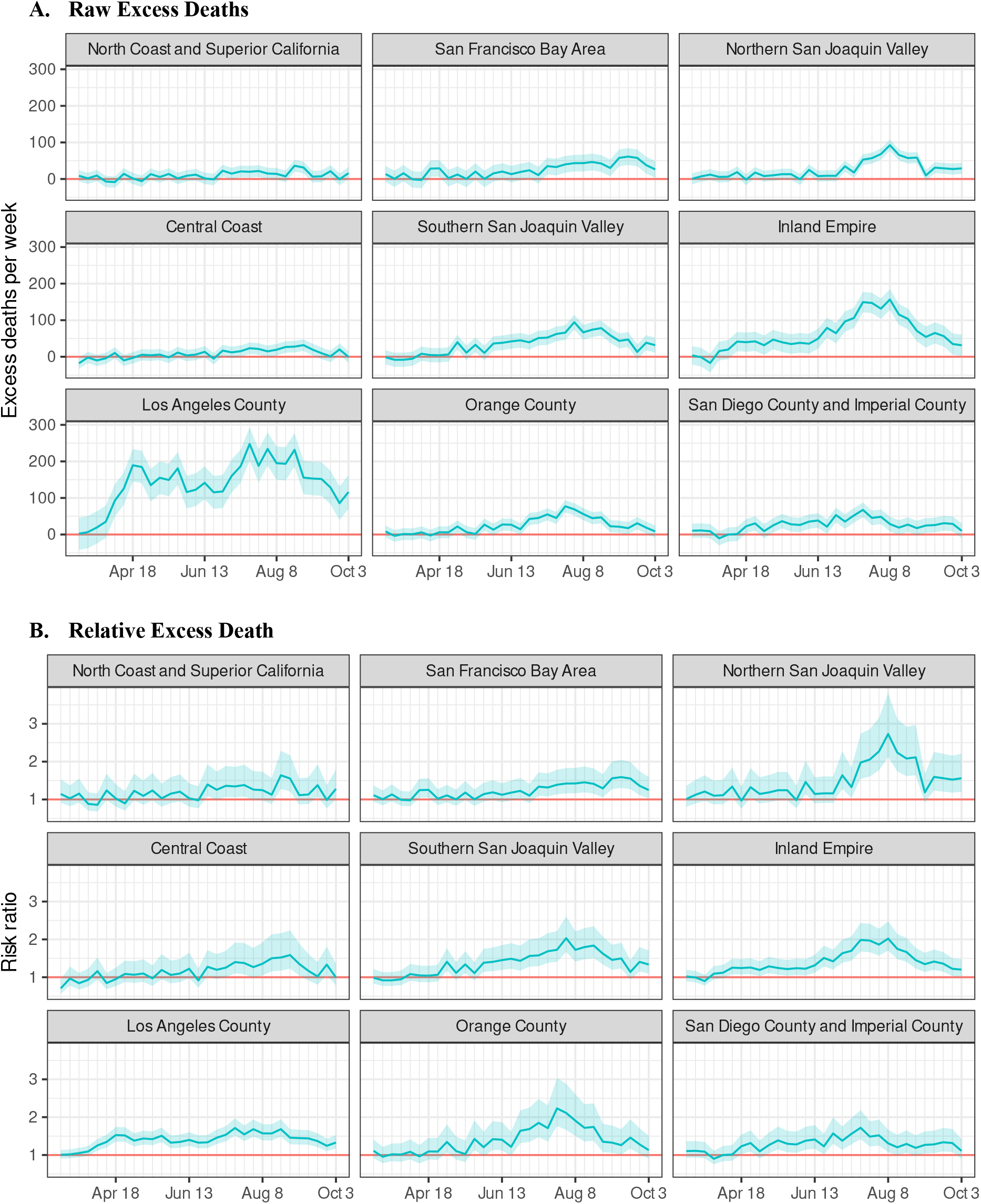
Weekly excess mortality attributable to the COVID-19 pandemic among Latinos in California, by region of death. * Risk ratio (expected/observed) for all-cause mortality, comparing the pandemic period (March to September 2020) to previous years. Source: California Department of Public Health Death Records.

**Figure S2.**
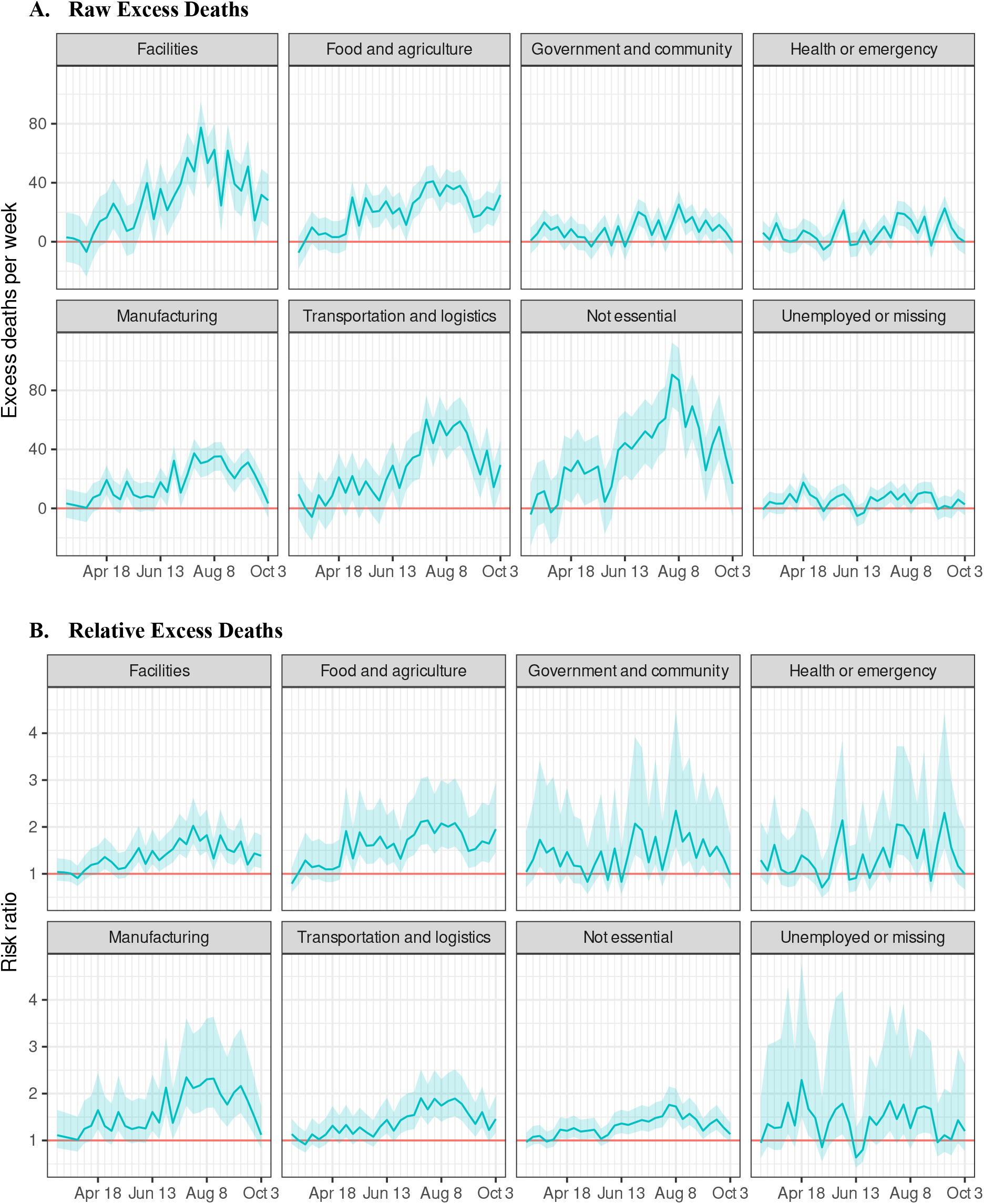
Weekly excess mortality attributable to the COVID-19 pandemic among Latinos in California, by occupational sector. * Risk ratio (expected/observed) for all-cause mortality, comparing the pandemic period (March to September 2020) to previous years. ** Includes only individuals ages 18 to 65 years of age. Source: California Department of Public Health Death Records.

**Figure S3.**
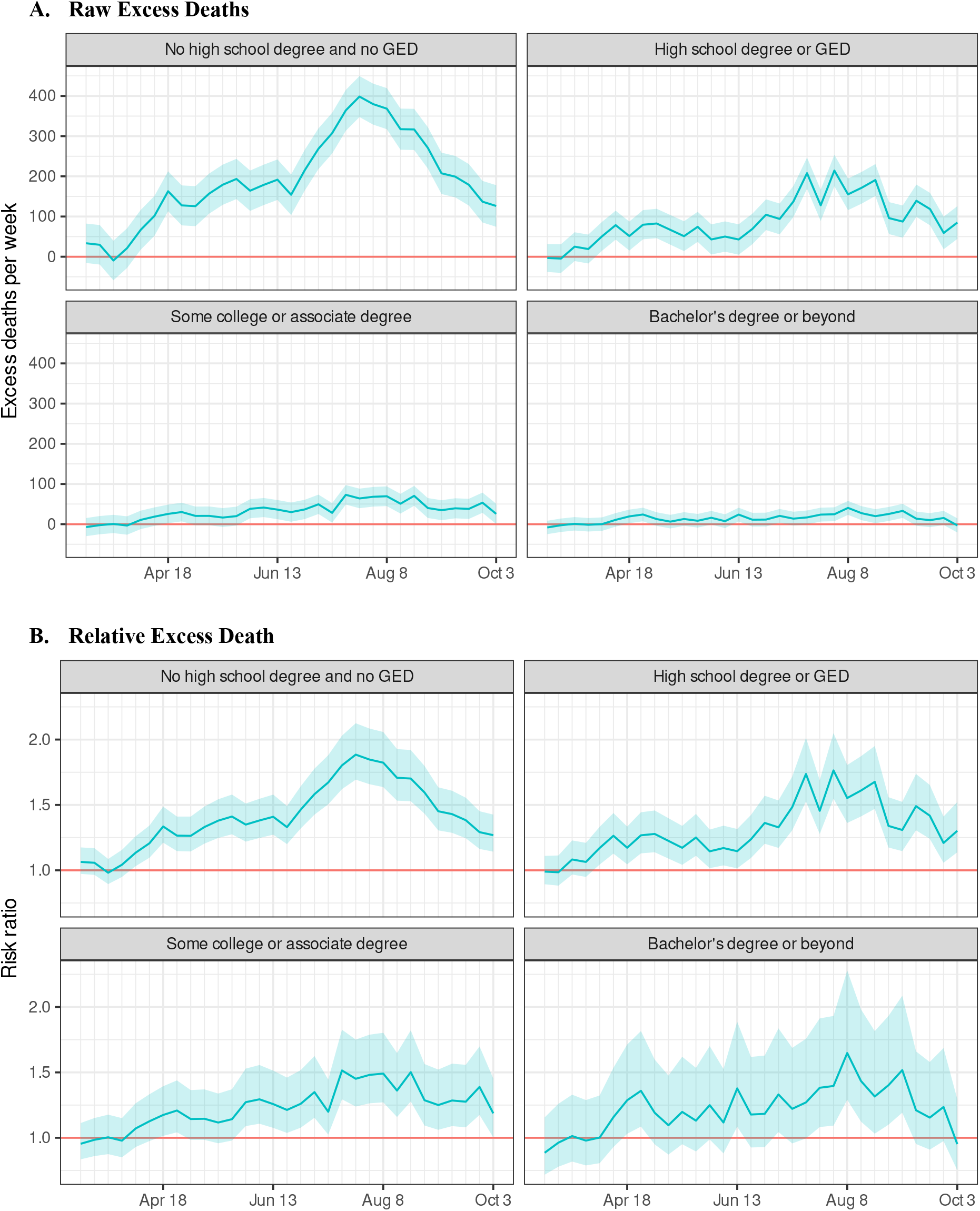
Weekly excess mortality attributable to the COVID-19 pandemic among Latinos in California, by educational attainment. * Risk ratio (expected/observed) for all-cause mortality, comparing the pandemic period (March to September 2020) to previous years. ** Includes only individuals ages 25 and older. Source: California Department of Public Health Death Records.

**Figure S4.**
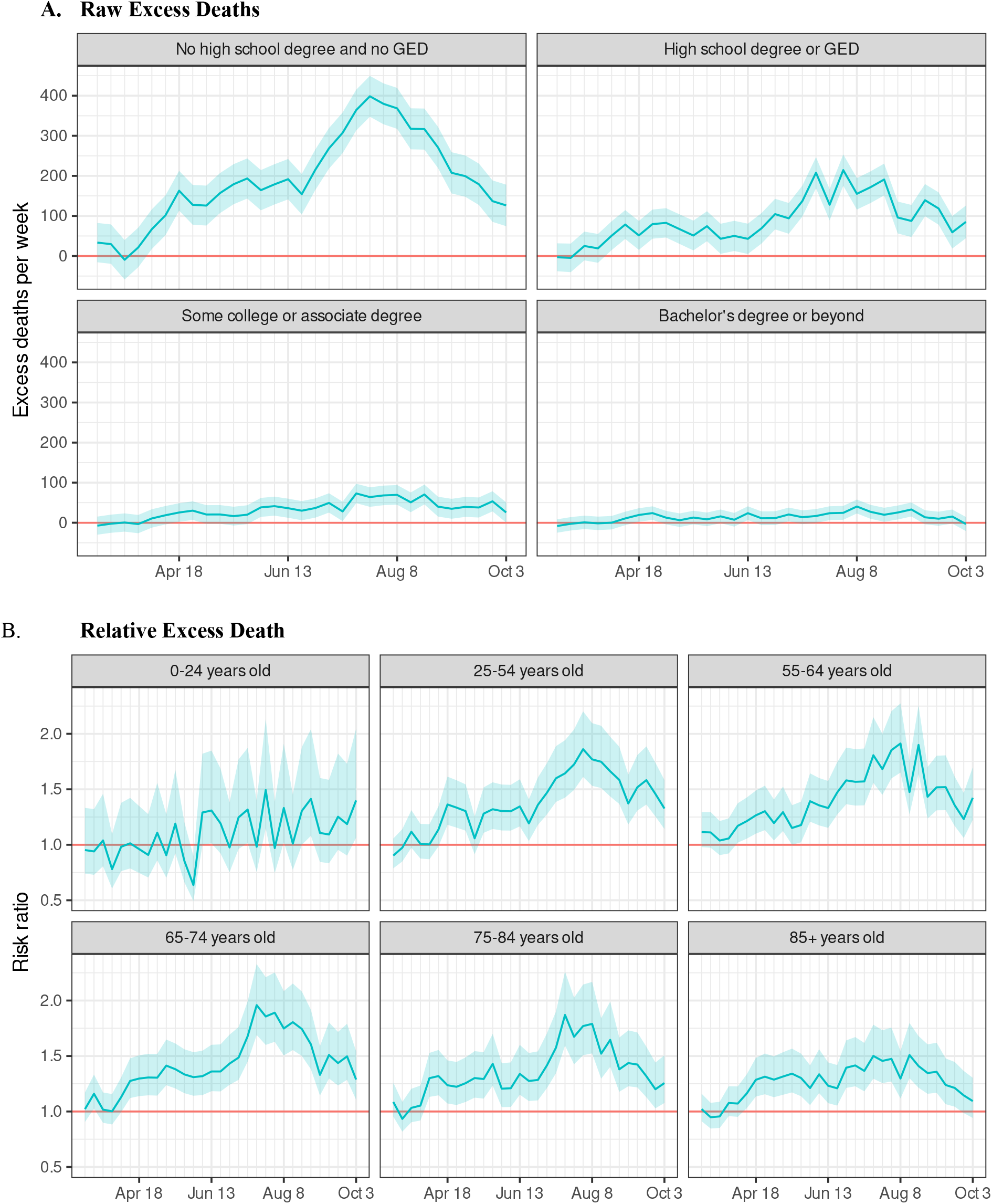
Weekly excess mortality attributable to the COVID-19 pandemic among Latinos in California, by age group. * Risk ratio (expected/observed) for all-cause mortality, comparing the pandemic period (March to September 2020) to previous years. Source: California Department of Public Health Death Records.

**Figure S5.**
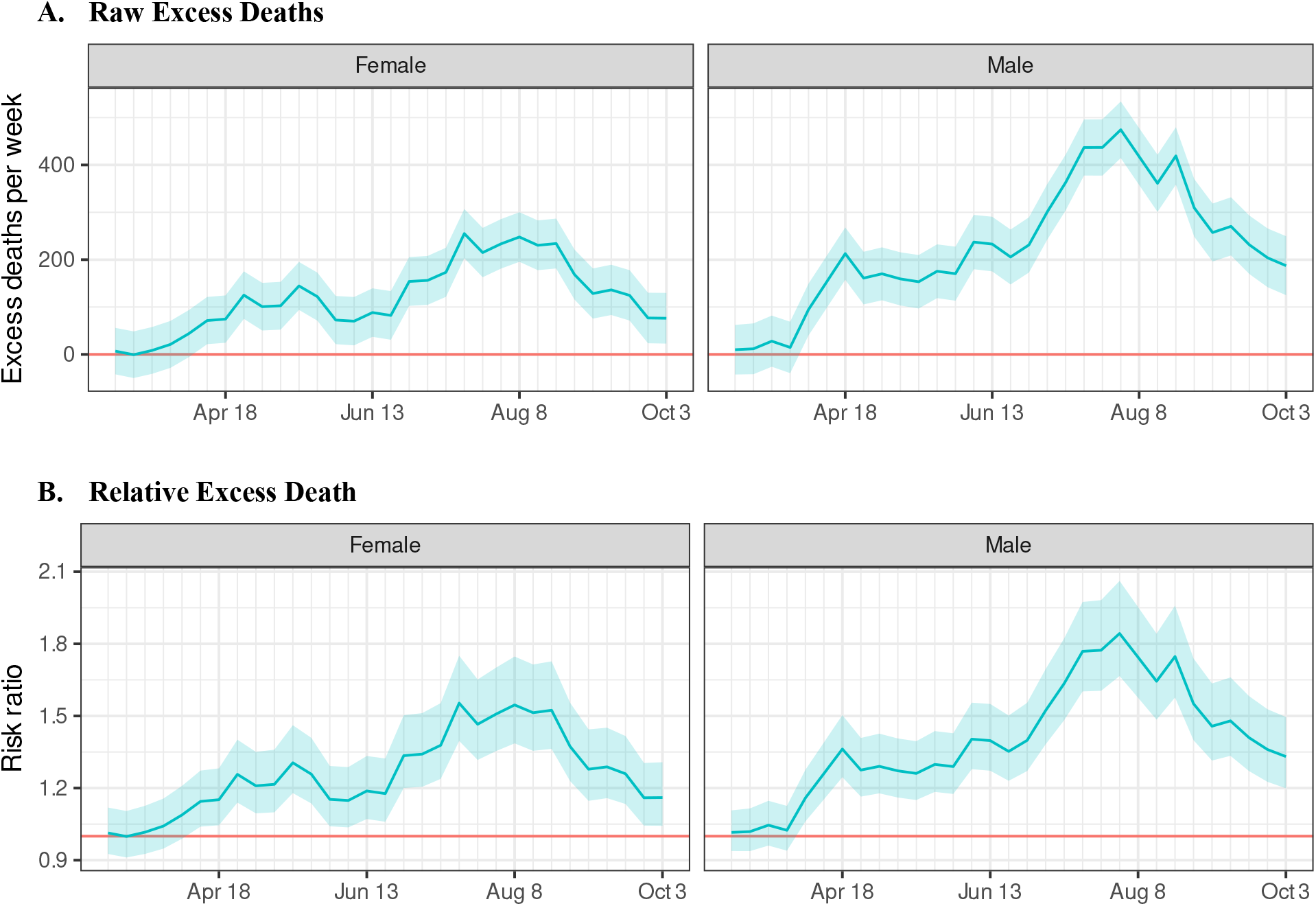
Weekly excess mortality attributable to the COVID-19 pandemic among Latinos in California, by sex. * Risk ratio (expected/observed) for all-cause mortality, comparing the pandemic period (March to September 2020) to previous years. Source: California Department of Public Health Death Records.

**Figure S6.**
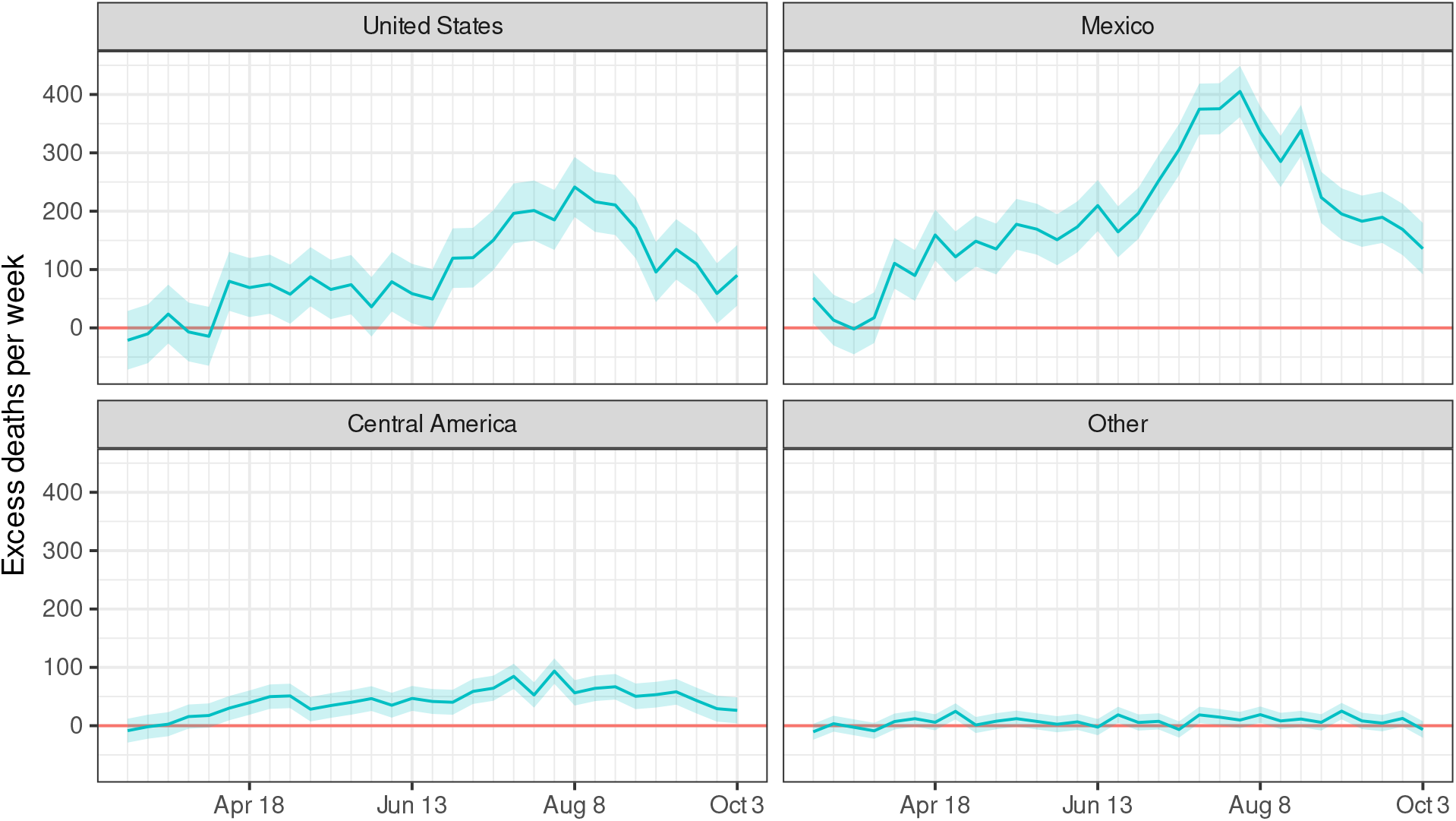
Raw weekly excess mortality attributable to the COVID-19 pandemic among Latinos in California, by place of birth. * Risk ratio (expected/observed) for all-cause mortality, comparing the pandemic period (March to September 2020) to previous years. Source: California Department of Public Health Death Records.

**Figure S7.**
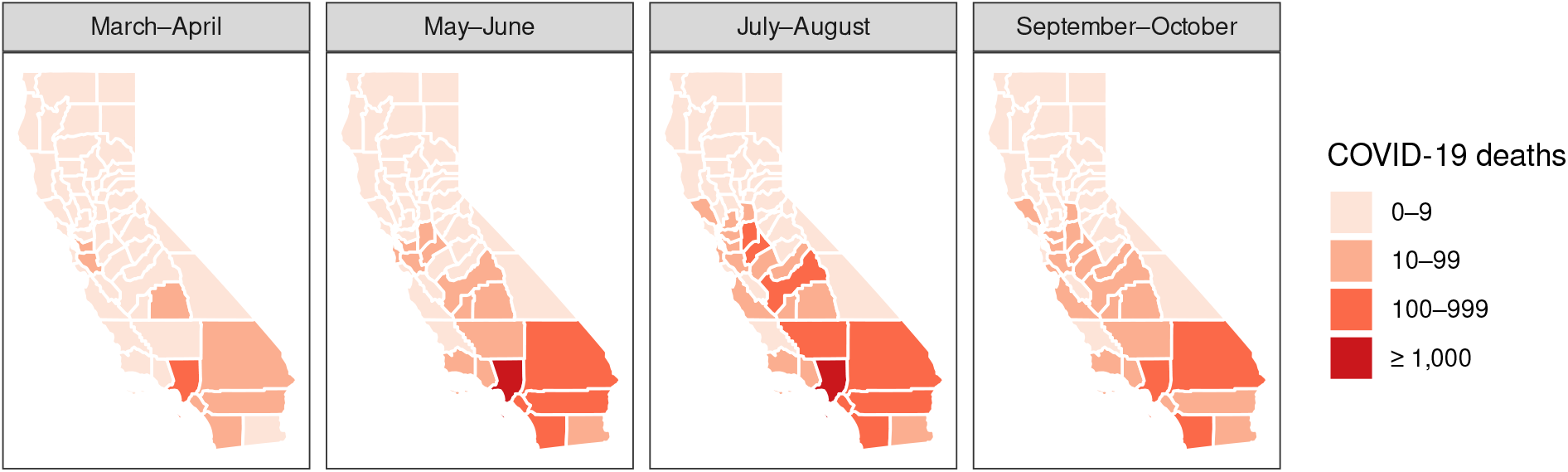
Map of COVID-19-confirmed mortality among Latinos in California, by period and region of death. Note: COVID-confirmed mortality among Latinos in California from March 1, 2020 to October 3, 2020, based on available death records as of December 28, 2020. Source: California Department of Public Health Death Records.

